# Investigating trends in those who experience menstrual bleeding changes after SARS-CoV-2 vaccination

**DOI:** 10.1101/2021.10.11.21264863

**Authors:** Katharine MN Lee, Eleanor J Junkins, Chongliang Luo, Urooba A Fatima, Maria L Cox, Kathryn BH Clancy

## Abstract

Early in 2021, many people began sharing that they experienced unexpected menstrual bleeding after SARS-CoV-2 inoculation. We investigated this emerging phenomenon of changed menstrual bleeding patterns among a convenience sample of currently and formerly menstruating people using a web-based survey. In this sample, 42% of people with regular menstrual cycles bled more heavily than usual while 44% reported no change after being vaccinated. Among respondents who typically do not menstruate, 71% of people on long-acting reversible contraceptives, 39% of people on gender-affirming hormones, and 66% of post-menopausal people reported breakthrough bleeding. We found increased/breakthrough bleeding was significantly associated with age, systemic vaccine side effects (fever, fatigue), history of pregnancy or birth, and ethnicity. Generally, changes to menstrual bleeding are not uncommon nor dangerous, yet attention to these experiences is necessary to build trust in medicine.

**Teaser:** Increased bleeding can occur post SARS-CoV-2 vaccines; this study investigates patterns in who experiences these changes.

## Introduction

Menstruating and formerly menstruating people began sharing that they experienced unexpected bleeding after being administered a SARS-CoV-2 vaccine in early 2021. Vaccine trial protocols do not typically monitor for major adverse events for more than seven days, and additional follow up communications do not inquire about menstrual cycles or bleeding.

Therefore, manufacturers had no way of addressing the extent to which this observation was a coincidence or a potential side effect of the vaccines. In media coverage, medical doctors and public health experts hastened to say that there was “no biological mechanism” or “no data” to support a relationship between vaccine administration and menstrual changes. In other cases experts declared that these changes were more likely a result of “stress” (*1–4*).

Unfortunately, dismissal by medical experts fueled greater concerns, as both vaccine hesitant and anti-vaccine individuals and organizations conflated the possibility of short-term menstrual changes with long-term harms to fertility. Pundits, politicians, religious leaders, and wellness influencers worked the oft-used framing of protecting women to advise against the vaccine (*5–9*). As the SARS-CoV-2 vaccine became available to adolescents, calls to understand the menstrual changes associated with the vaccine increased as parents felt they were weighing their child’s pubertal development and future fertility against their risk of becoming sick with COVID-19 (*10, 11*).

There are in fact multiple plausible biological mechanisms to explain a relationship between an acute immune challenge like a vaccine (*12*), its corresponding and well-known systemic effects on hemostasis and inflammation (*13*), and menstrual repair mechanisms of the uterus (*14–17*). The uterine reproductive system is flexible and adaptable in the face of stressors, in order to weather short-term challenges in a way that leaves long-term fertility intact (*18, 19*). We know that running a marathon may influence hormone concentrations in the short term while not rendering that person infertile (*20*); that short-term calorie restriction that results in a loss of menstrual cycling can be overcome by resuming normal feeding (*21*); that inflammation influences ovarian hormones (*22–24*); and that psychosocial stressors can correspond to cycle irregularity and yet resilience can buffer one from these harms (*25–27*). Less severe, short-term stressors can and do influence menstrual cycling and menstruation, and this has been established over forty years of cycle research (*19, 20, 28–30*). This work has also established that while sustained early stressors can influence adult hormone concentrations, short-term stressors resolve and do not produce long-term effects (*31*). The immune response invoked by a vaccine is quite different from the sustained immune assault of COVID-19 itself: studies and anecdotal reports are already demonstrating that menstrual function may be disrupted long-term, particularly in those with Long COVID (*32–35*).

Vaccines function by mobilizing the immune system to protect from disease if exposure occurs. This immune activation is important, although it may also produce a cascade of other localized (e.g., soreness at injection site) or systemic (e.g., fatigue, fever) inflammatory responses. Studies that assess the direct effect of vaccination on the menstrual cycle are few and far between. A study from 1913 identified that the typhoid vaccine was associated with menstrual irregularities, which included missed, late, and early menstruation, discomfort, and heavy bleeding in more than half of their female sample (*36*). Hepatitis B studies have also indicated that menstruation could be altered (*37*), and a HPV post-market safety study found that over a quarter of participants reported menstrual irregularity (*38*) though ovarian insufficiency, a type of reduced fertility analyzed because of case reports, is not associated with this vaccine (*39*). The speed and coverage of the current COVID-19 pandemic and vaccination campaign may have inadvertently highlighted a previously under-recognized side effect of especially immunogenic vaccines administered in adulthood, which is that systemic inflammatory responses may in some individuals invoke downstream responses in target organs such as the uterus.

The question of whether and when the particular acute immune challenge of the current SARS-CoV-2 vaccines affects menstrual cycling or menstruation is an emerging one with limitations on study design. Given the vaccines’ overall established safety generally (*40–42*) and in relation to fertility and pregnancy (*43–47*), and the multiple waves of viral spread and variant emergence the world has endured with this deadly pandemic, we opted for an observational and retrospective study design of vaccinated people rather than a prospective design with a control or crossover group of unvaccinated individuals. In early anecdotal reports of menstrual cycle experiences, the nature and breadth of the cycle changes were unclear: among those experiencing side effects were people experiencing earlier, later, heavier, lighter periods? Were other menstrual cycle phenomena also altered, like midcycle and premenstrual experiences? Were formerly menstruating people (e.g., those on menstrual suppression therapies or postmenopausal people) affected?

For this reason, we established an emergent, exploratory, mixed methods survey instrument intended to capture a wide range of responses from current and formerly menstruating adults. Here we share results from our first round of analyses (N=39,129), as well as the ways that this early exploration has made it possible to establish the parameters of the phenomenon of post-vaccine menstrual change. We focus on findings related to menstrual bleeding (in people who menstruate regularly) or breakthrough bleeding (in people who do not currently menstruate) from the first three months of data collection in order to provide a description of trends to clinicians and the public alike. Specifically, we sought to address the following research questions: 1) What is the range of menstrual bleeding changes reported by regularly menstruating respondents after being administered the SARS-CoV-2 vaccine? 2) To what extent are non-menstruating respondents reporting breakthrough bleeding after being administered the SARS-CoV-2 vaccine? And 3) Are there trends among those with a changed bleeding pattern to help determine proximate mechanisms acting on the uterus?

Answers derived from this convenience observational sample can help shape the narrative around the nature of short-term menstrual changes, help clinicians working with vaccine hesitant patients, and develop the necessary, on-the-ground data on this new phenomenon to design future prospective, mechanistic studies on the relationship between vaccine immune responses and menstrual repair. Projects that take the time to establish trends and listen to respondents are important first steps to understanding details of emerging health concerns (*48*).

## Results

### Demographics and summary statistics

After data cleaning and aggregation of the first three months of data (Fig. 1), respondents in our sample (N=39,129) were between 18 to 80 years old (median=33 years; *M*_age_=34.22 years, SD=9.18). All participants were fully vaccinated (at least fourteen days after one or two required doses as this was before boosters) and had not contracted COVID-19 (diagnosed or suspected). This sample included 35,572 (90.9%) woman-only identifying and 3,557 (9.1%) gender diverse respondents; 32,983 (84.3%) white-only identifying and 6,146 (15.7%) racially diverse respondents; and 31,134 (79.6%) non-Hispanic or Latinx and 7,995 (20.4%) Hispanic, Latinx, or other respondents (summary demographics in Table 1; more detailed in Table S1).

**Fig. 1.**
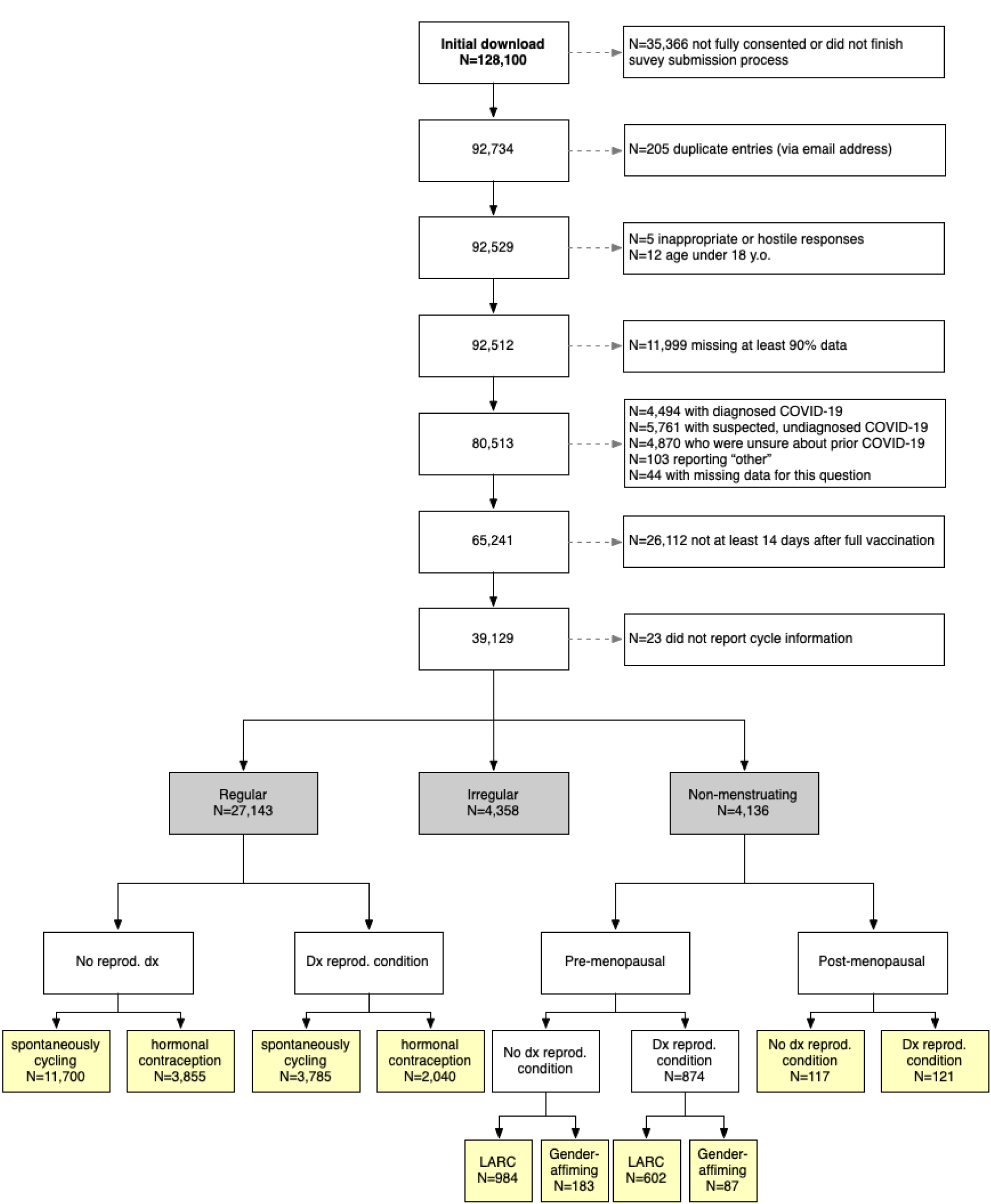
Flowchart of data cleaning and aggregation. Note that totals in the yellow boxes do not add up to the numbers in the grey boxes due to uncertain menopause stage (n=1,522), currently or recently lactating (n=2,498), having had a hysterectomy (n=43), discrepant responses (e.g., self-reported period details did not align with self-reported menstrual group), and the divisions made to the samples. Further details can be found in Supplemental Materials.

**Table 1.**
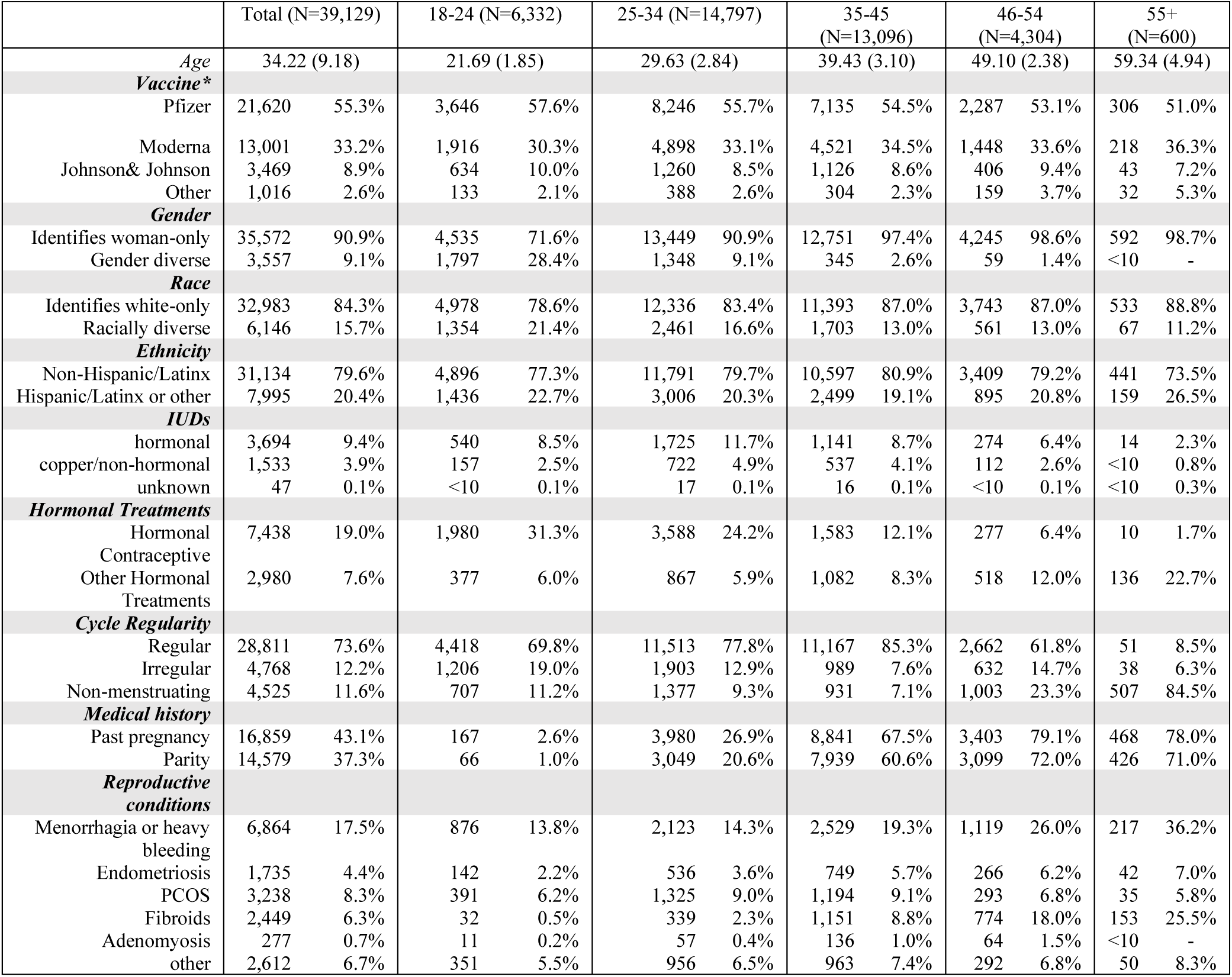
Sample summary information. Demographics and sample background in life stages corresponding to later sample restrictions (pre-menopausal 18-45, menopause transition or perimenopause 46-54, post-menopause 55+). Note: <10 was used for any cells with fewer than 10 individuals. Ages are binned based on approximate life stages and the sample restrictions.

Respondents were vaccinated with Pfizer (N=21,620), Moderna, (N=13,001), AstraZeneca (N=751), Johnson & Johnson (N=3,469), Novavax (N=61), or other (N=204) vaccines, with 23 not reporting vaccine type. Self-report of localized vaccine side effects (soreness at injection site) after the first dose and second dose were 87.6% and 77.4%, respectively, when combined across all vaccine types. After the first and second dose, 54.3% and 74.6% of respondents (respectively) report experiencing at least one of the common systemic vaccine side effects (headache, nausea, fever, and/or fatigue). Of those that reported systemic vaccine side effects, 40.6% experienced systemic effects after both doses. Vaccine symptoms, period flow changes, period symptoms, and timing of period symptoms reported by study respondents are presented by age categories (Fig. 2, detailed reporting by vaccine type in Table S2). The Johnson & Johnson vaccine, being the only single dose vaccine at the time of survey, was excluded from later analyses.

**Fig. 2.**
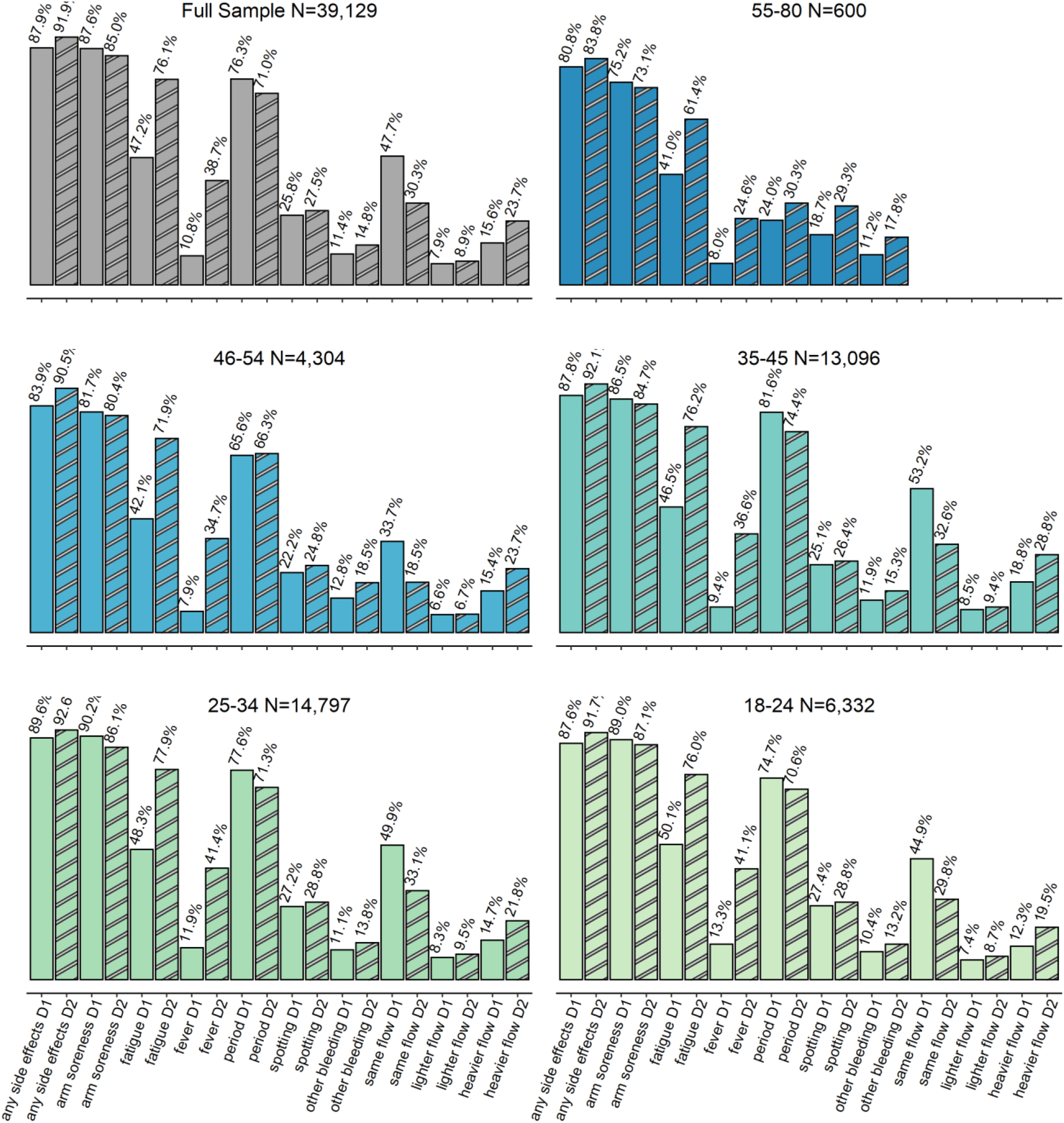
Descriptive statistics of the full sample (dose 1 displayed in solid and dose 2 displayed striped). The most salient vaccine and menstrual side effects pertaining to the analysis are presented here. The sample sizes of dose 2 variables decrease because of those who received one-dose Johnson & Johnson vaccine. The respective samples become: full N=35,660, 18-24 N=5,698, 25-34 N=13,537, 35-45 N=11,970, 46-54 N=3,898, 55-80 N=557.

### Reported menstrual changes in regularly menstruating people

Respondents reported noticing changes to their period 1-7 days after vaccines (Dose 1: 31.4%; Dose 2: 37.0%), 8-14 days after vaccines (Dose 1: 25.9%; Dose 2: 23.6%), or more than 14 after receiving their vaccines (Dose 1: 29.9%; Dose 2: 26.8%), with the rest of respondents reporting they were menstruating when they received the vaccine (Dose1: 12.7%; Dose 2: 12.5%). In total, 42.1% reported heavier menstrual flow after vaccines, 14.3% reported not heavier (characterized by a mix of lighter or no change) menstrual flow, and 43.6% reported no change to flow after vaccines.

#### Associations with a heavier post-vaccine menstrual flow

Following the univariate tests for association (Table S6), we fit a multivariate logistic regression model to explore the relationship between heavier menstrual bleeding after vaccination and several factors: vaccine type, demographic factors, reproductive history, hormonal contraception use, and systemic vaccine response (Fig. 3a). Our main findings were that a heavier menstrual flow was more likely for those respondents who were: non-white race, Hispanic/Latinx, older, had a diagnosed reproductive condition, used hormonal contraception, had been pregnant in the past (whether or not they had given birth), were parous, or experienced fever or fatigue after vaccination. The comparison between those who have given birth and those who have not is conditioned on having been pregnant: the combination of having been pregnant but not given birth is associated with the highest risk of heavier flow. We note that vaccine type, race, and use of hormonal contraceptives have odds ratios and 95% confidence intervals very close to or overlapping with 1 in combination with a relatively high p-value, suggesting they have negligible or relatively small effects in this model.

**Fig. 3.**
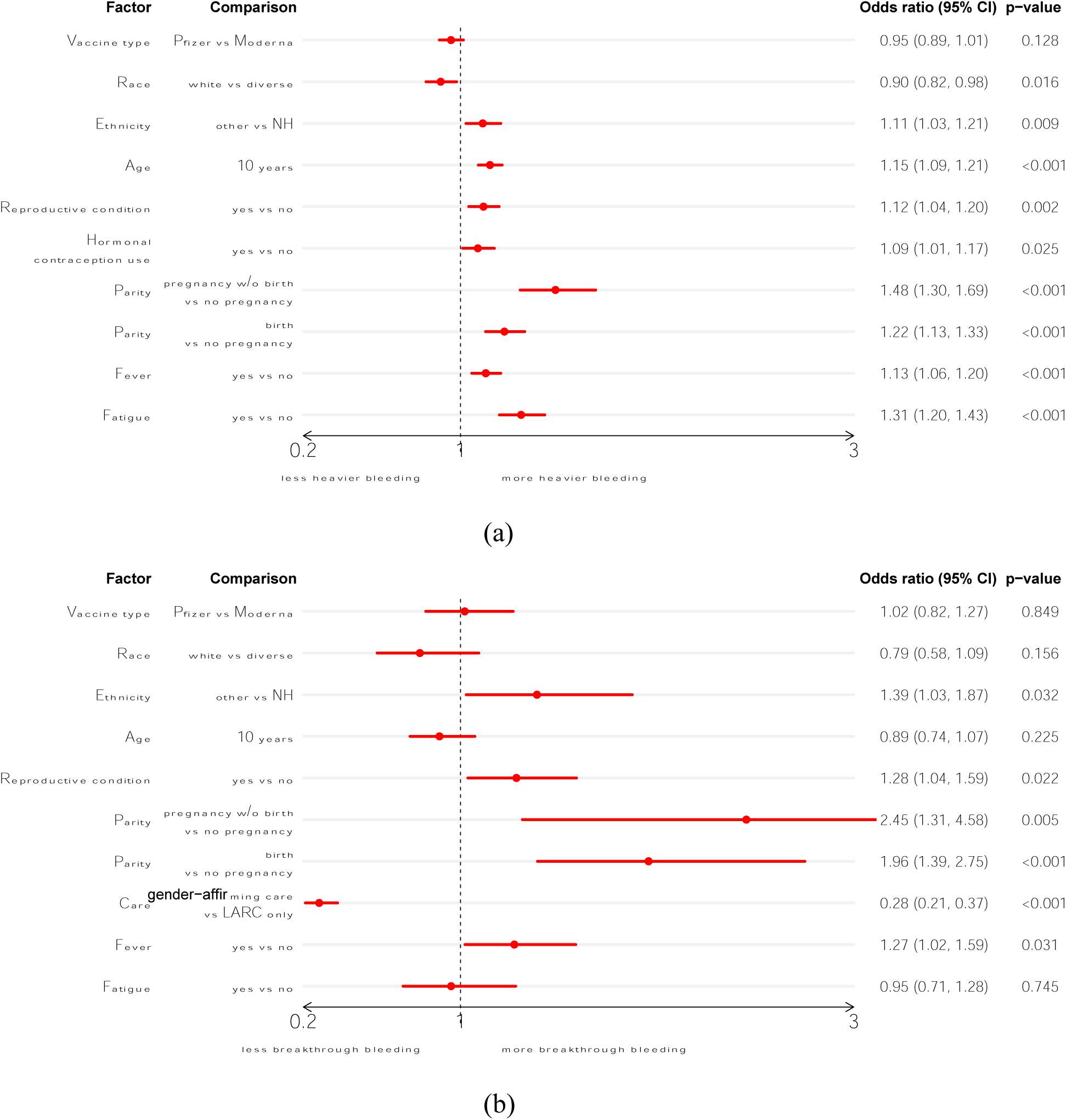
Multivariate logistic regression of (a) heavier flow in the regularly menstruating group (N=17,113, after removing those respondents with vaccines other than Pfizer or Moderna, or missing parity history or flow change); and (b) breakthrough bleeding in the non-menstruating premenopausal group (N=1,771, after removing those respondents with vaccines other than Pfizer or Moderna, or missing parity history) after either dose of the vaccine. The graph presents the ratio of the odds of heavy bleeding occurring in the first group of the comparison vs. the second group (except for Age, which is in 10-year increments). If the odds ratio is greater than 1, the first group in the comparison has higher risk of experiencing heavier bleeding (or breakthrough bleeding). NH=Not Hispanic/LatinX.

#### Reproductive conditions

We additionally examined the relationship of specific reproductive conditions often associated with altered menstrual bleeding by comparing respondents with diagnosed conditions to respondents with no reported reproductive conditions (Fig. 4). A higher proportion of respondents with endometriosis (51.1%), menorrhagia (44.3%), fibroids (49.1%), PCOS (46.2%), and adenomyosis (54.9%) reported experiencing a heavier menstrual flow post-vaccine than the respondents without diagnosed reproductive conditions (40.9%). Odds ratios and chi-squared results for these groups are in Table S7.

**Fig. 4.**
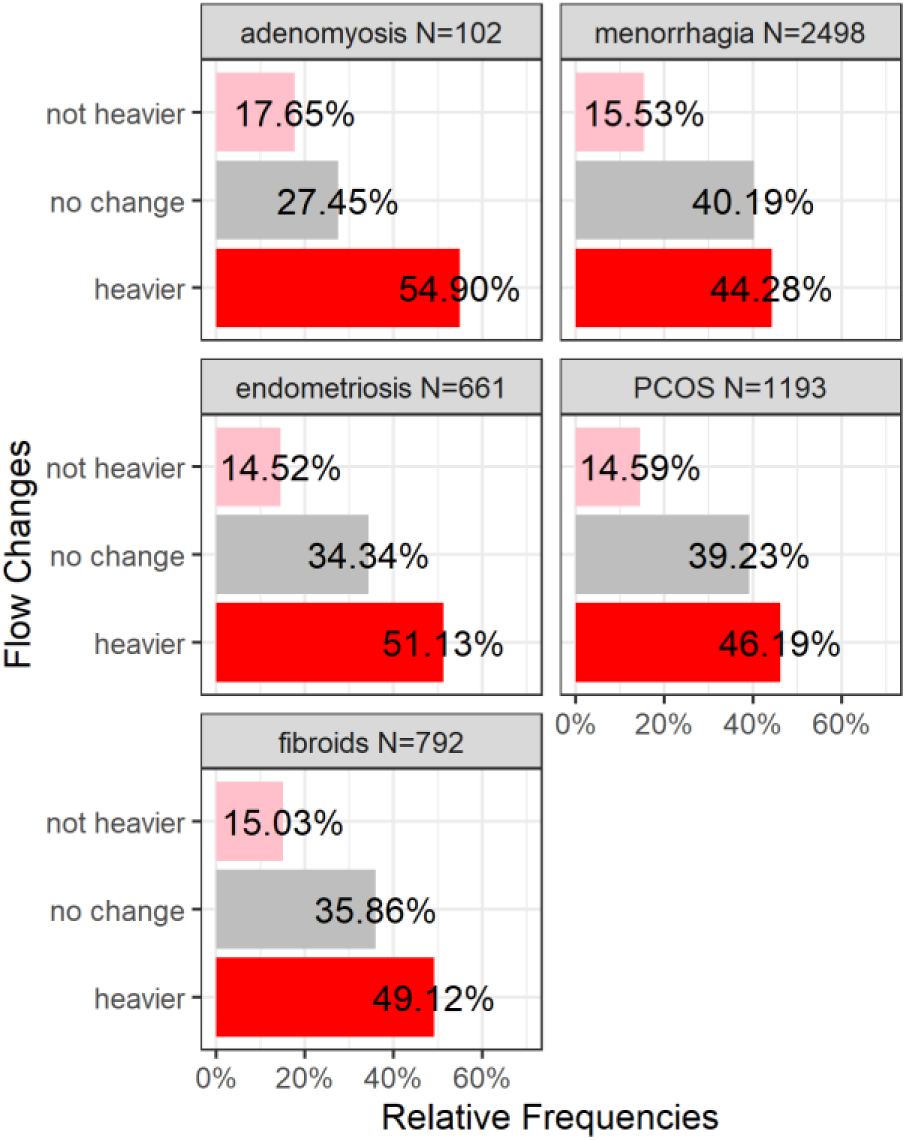
Menstrual flow changes in regularly cycling individuals with diagnosed reproductive conditions. Displayed on the x-axis are the percentage of individuals reporting each flow change condition (y-axis).

### Reported breakthrough bleeding in non-menstruating respondents

Non-menstruating people consisted of two groups: premenopausal people (using LARC and/or continuous hormonal contraceptives and/or gender-affirming treatment that eliminates menstruation) and postmenopausal people over the age of 55 who had not bled for at least 12 months (prior to SARS-CoV-2 vaccination). Among non-menstruating, premenopausal respondents (N=1,815) on hormonal treatments, a majority (65.7%) experienced breakthrough bleeding after a vaccine, although this was significantly different between respondents using only LARC (70.5%) and respondents using gender-affirming care (38.5%). Among post-menopausal people who were not on any hormonal treatments (N=238) breakthrough bleeding was reported by 66.0% of respondents (Fig. 5).

**Fig. 5.**
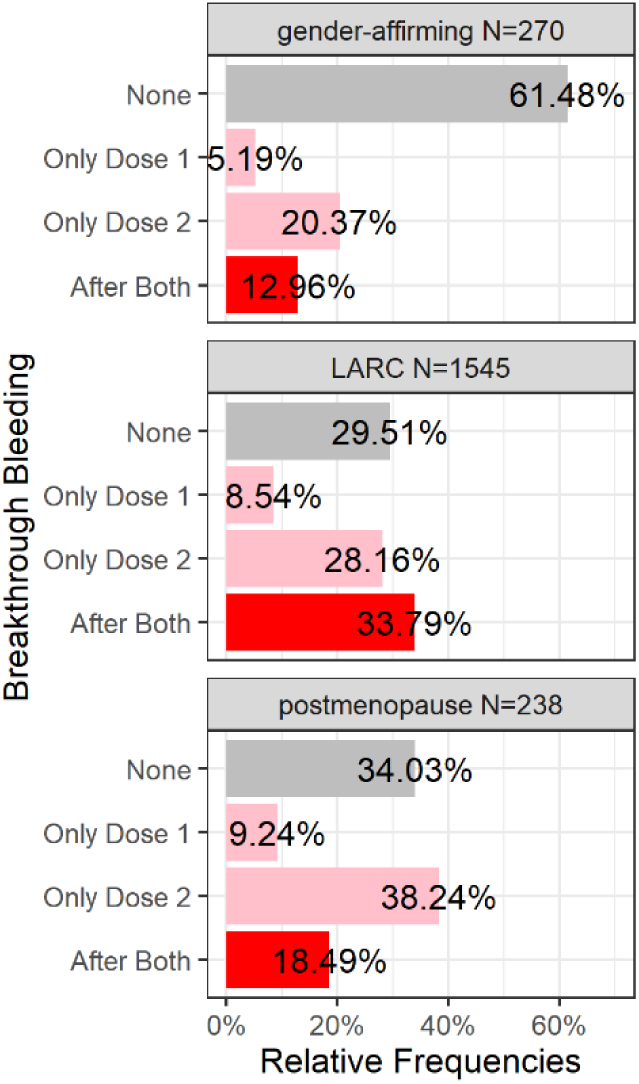
Breakthrough bleeding in non-menstruating individuals. Displayed on the x-axis are the percentage of individuals reporting breakthrough bleeding after both doses, only following dose 2, only following dose 1, or no breakthrough bleeding during vaccination time (y-axis).

**Fig 6.**
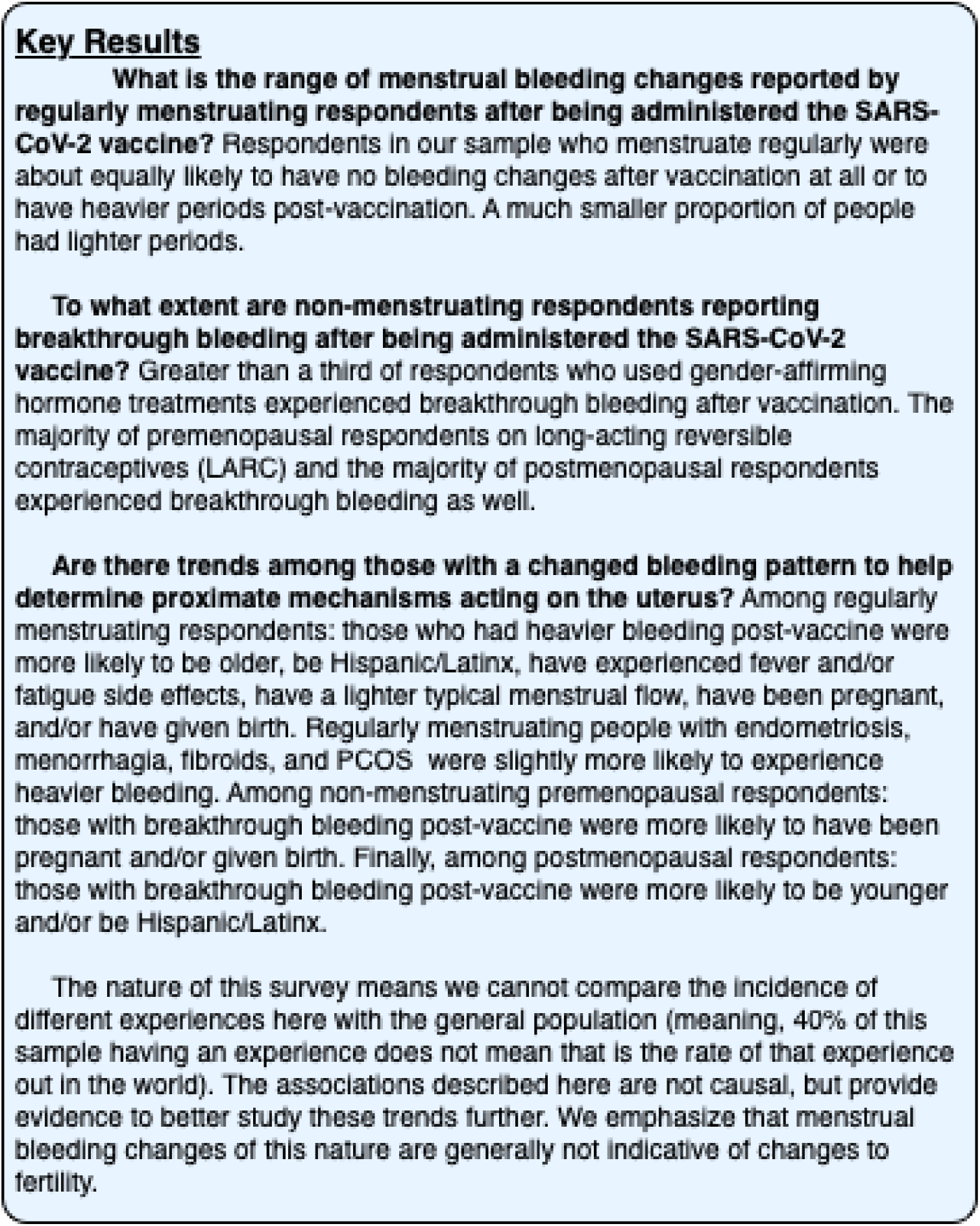
Summary of key results.

#### Associations with breakthrough bleeding in premenopausal respondents

A multivariate logistic regression of breakthrough bleeding in the non-menstruating premenopausal group after either dose of the vaccine was fitted. The result (Fig. 3b) shows an increased chance of breakthrough bleeding for those respondents who were: Hispanic/Latinx, had been pregnant in the past but had not given birth, had a diagnosed reproductive condition, LARC only, or experienced fever after vaccination.

#### Associations with breakthrough bleeding in post-menopausal respondents

Age was significantly different between those that experienced breakthrough bleeding occurrence or not (*t*(147.99) = -2.255, *p*=0.026), with post-menopausal people who experienced breakthrough bleeding being slightly younger (*M*=59.8 years) than those who did not (*M=*61.4 years). Ethnicity was associated with breakthrough bleeding, with non-Hispanic/Latinx respondents being less likely to report breakthrough bleeding. There was no significant difference in rate of occurrence of breakthrough bleeding by vaccine type, systemic side effects of fever or fatigue, or reproductive history of past pregnancy or parity (Table 2).

**Table 2.**
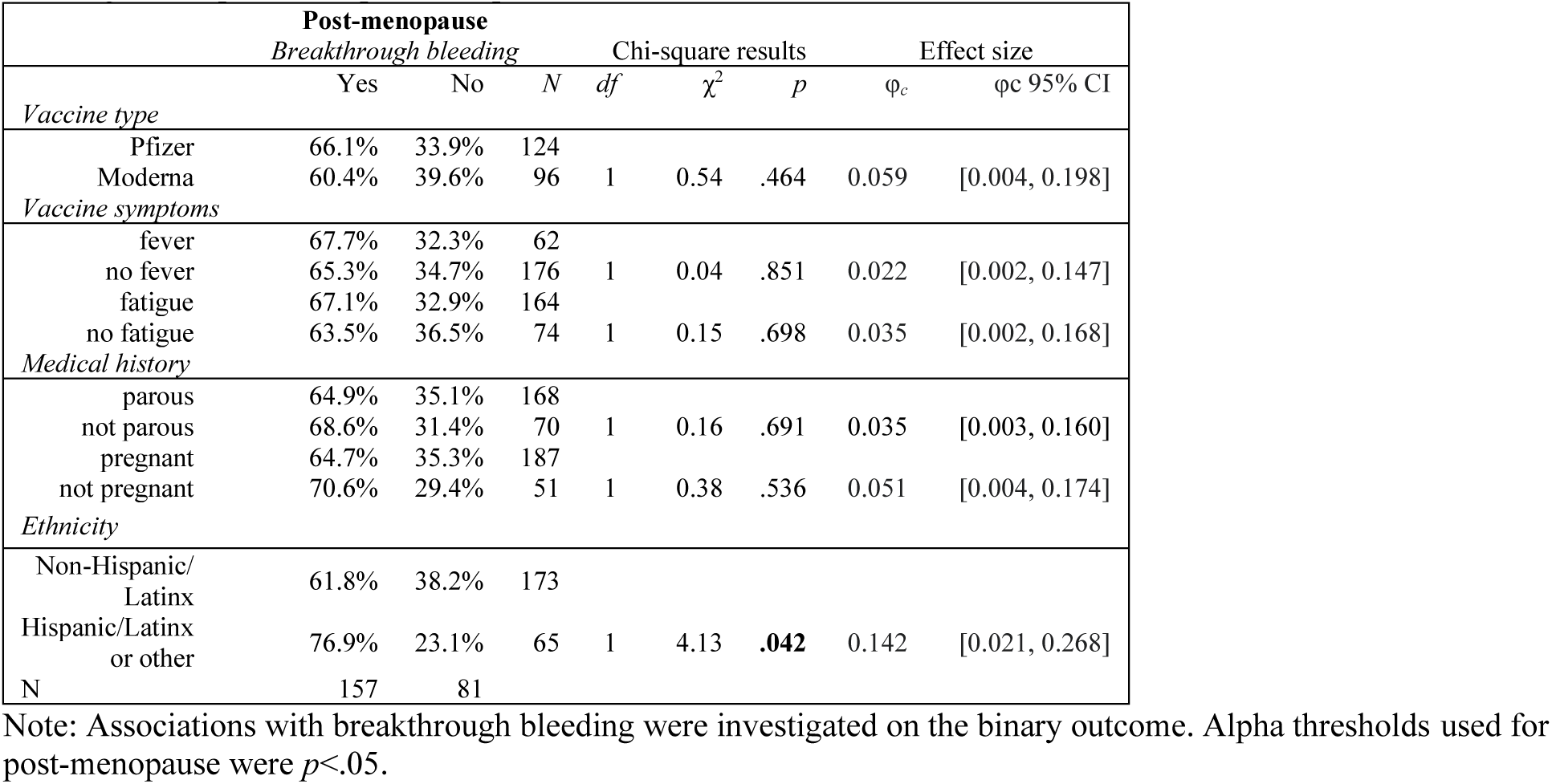
Breakthrough bleeding in post-menopausal people. Vaccine and medical history related to breakthrough bleeding across post-menopausal respondents.

## Discussion

We present initial summary statistics and descriptive analyses of changes to menstrual bleeding in a large and diverse sample of currently and formerly menstruating adults after SARS-CoV-2 vaccination. This is the very first characterization of post-vaccine menstrual bleeding changes for a gender-diverse sample of pre- and post-menopausal people. We cannot estimate prevalence or incidence based on our methodological approach of this emergent phenomenon, and the associations reported here cannot establish causality. However, our results highlight that many people noted changes in their menstrual bleeding, with 56% of our regularly menstruating sample reporting some change in their menstrual bleeding after one or more of the vaccine doses. Furthermore some of the trends we observe support hypothesis development for additional prospective studies in hemostatic and inflammatory changes to the endometrium after an acute immune response (Fig. 4).

In this first analysis, we focus on the heavier bleeding of currently menstruating and breakthrough bleeding of formerly menstruating people, which we define as an increased bleeding phenotype. The increased bleeding phenotype appeared to be the most common post-vaccination change within our sample. Initial forays into our qualitative data suggest a widely variable experience of the increased bleeding phenotype, confounding a straightforward case definition. At this time, we suggest that rather than a threshold quantity to define the increased bleeding phenotype, vaccinated people and providers instead consider menstrual changes in the context of what is typical for the vaccinated person. This definition is in line with recent changes to how heavy menstrual bleeding is described clinically (*49*), focusing more on lived experience and quality of life change than a particular quantity of blood loss.

Increased bleeding is often distressing, and it can (and often should) lead providers towards diagnostic procedures to assess its origins (*50–52*). This is especially true when it comes to breakthrough bleeding among formerly menstruating people, for whom this symptom can be an early sign of cancer. When possible side effects to a medical treatment are not shared with the clinical or patient population, it may lead to unnecessary, painful, and expensive diagnostic procedures. For example, several studies have now shown that epidurals likely increase the risk of heavy and breakthrough bleeding among regularly cycling and postmenopausal people, respectively (*53, 54*). In one recent study, 17% of postmenopausal respondents reported breakthrough bleeding after injection, versus 7% from a control group. Of the thirty-one respondents who reported this bleeding to their physician, thirteen had endometrial biopsies collected, and two had transvaginal ultrasounds. While all results were benign, endometrial biopsies are known to be painful and invasive procedures (*55, 56*). Though these data have been reported in the literature for at least a decade, no patient-facing information about epidurals that we could find makes note of the risk of unexpected bleeding, which means potentially unnecessary, expensive, painful diagnostic procedures may continue today.

Unexpected bleeding has other major and even life-threatening consequences. Trans men, trans masculine people, and masculine of center genderqueer people, many of whom suppress periods with a combination of LARC and masculinizing therapies, may find themselves suddenly navigating public bathrooms or workplaces while menstruating. Therefore, this unexpected bleeding runs the risk of psychological distress for those who experience gender dysphoria with menstruation and physical harm for people for whom managing menstruation in public is dangerous (*57, 58*).

In addition to our finding of a significant proportion of respondents experiencing some form of increased bleeding, we noticed some trends in who was more likely to have this phenotype. Among premenopausal 18–45-year-old respondents, those who were older and/or Hispanic or Latinx (using U.S. census demographic approaches) were more likely to report heavier bleeding post-vaccine. Prior pregnancy and prior birth also were associated with a greater risk of heavier bleeding. Finally, premenopausal menstruating respondents who were diagnosed with endometriosis, menorrhagia, fibroids, adenomyosis, and/or PCOS were more likely to report experiencing heavier bleeding post-vaccine compared to those without any diagnosed reproductive condition. We also find that many respondents who had post-vaccine changes did not have them until more than a week post-inoculation, which extends beyond the typical seven days of adverse symptom reporting in vaccine trials.

The responsiveness of menstrual cycles and bleeding patterns to external stressors is well known (*59*). Responsiveness to external stressors is one reason menstrual cycles are often thought of as reflecting overall health status, or a so-called “vital sign” in clinical practice (*60–62*). Thus, many people are attuned to menstrual cycles and take note of changes as potentially indicating other underlying health concerns. For many people, menstruation matters for reasons beyond current conceptive intentions: menstruation relates to their experiences of gender and gender dysphoria, to their intuitive connections to bodily processes, and to their fears and embarrassments surrounding menstrual stigma (*57, 63, 64*). Therefore, unexpected and unplanned menstrual changes can cause concern, distress, or other negative responses, in addition to discomfort and physical pain.

Despite this, menstruation is seldom considered a variable during vaccine trials aside from determining last menstrual period as part of established protections against volunteers being or getting pregnant. The vast majority of research that has been conducted regarding reproductive and menstrual function centers around whether or not live and attenuated vaccines are safe to give someone who is pregnant (*65–68*), or if it affects fertility (*69, 70*). The research that has been conducted on menstrual cycles specifically is often not able to establish a causal link, as the data is obtained through retrospective surveys or data mining (*71, 72*) and randomized controlled trials often do not allow a mechanism for reporting these changes (*73*). Data mining and signal detection in VAERS has resulted in the identification of several possible effects on menstruation that suggest further research is needed (*71, 72*); however, queries about changes in menstruation are still not a standard part of vaccine trials despite recent calls for more study (*74*).

Menstruation is an inflammatory and hemorrhagic event that must be resolved quickly to restore uterine function and prevent infection and continued hemorrhage (*14, 75*). Disruption of the normal coagulation pathway of the endometrium may delay the repair mechanisms that allow menses to end quickly. A few of our findings suggest that vaccination is less likely to be affecting periods via ovarian hormone pathways, and more likely along these inflammatory pathways. For instance, we found little difference between respondents with spontaneous and hormonally contracepting cycles in the rate of post-vaccine heavy menstrual flow. If changes in menstrual bleeding were due to vaccine-related disruption of ovarian hormones, we would expect that regularly menstruating people taking hormonal contraception would be far less likely to experience changes, as their cycles are largely regulated by exogenous hormones. We also found a significant proportion of formerly menstruating people, including post-menopausal participants with presumably dormant ovaries, experienced breakthrough bleeding. The greater presence of this increased bleeding phenotype among regularly cycling premenopausal respondents who were older and/or parous points to ways in which mature and established menstrual repair mechanisms may create a vulnerability to this short-term phenomenon. In addition, the greater proportion of people with certain reproductive conditions experiencing heavier bleeding post-vaccine could also point to a vulnerability among those with hyperproliferative and/or vascular/hemostatic conditions.

Data used in these analyses are unable to represent population prevalence of post-vaccine menstrual changes. They may be biased towards those who noted some change in their own menstrual or bleeding experiences, particularly if that change was uncomfortable, painful, frightening, or concerning. That said, a significant portion of the respondents who took part reported no menstrual change. Evidence suggests that people with other types of negative experiences are less, not more likely to participate in surveys where they suspect they will be expected to recount such material (*76, 77*). A large number of qualitative responses alluded to the fact that people who are interested in science or cared about the research participated despite not having adverse menstrual experiences post-vaccine.

Respondents in our sample were more likely to report fever (which was associated with heavier bleeding in our analyses) than participants in published vaccine safety and efficacy studies. Percentage comparisons between vaccines and studies, however, can be complicated by several factors which include the age distribution of the sample, size of the sample, how the data is collected, and how the factor is defined to participants. Studies of multiple SARS-CoV-2 vaccine types indicated that younger participants reported higher incidence of fever than older participants (*40, 41, 78–81*). Vaccine safety studies that use a self-report measure of “feeling feverish” report higher percentages of affected participants compared to those that measured temperature (*79, 81, 82*). Based on literature across vaccine trial results, between 0.8% -17.4% of participants reported having a fever after vaccination regardless of dose number, and between 2.5% -71% experienced fatigue (*40, 41, 78–83*). Our survey asked participants to self-report fever, and so we may have a significant portion of the sample who experienced elevated temperature that did not meet the clinical criterion. The other possibility is that those more likely to experience menstrual change are more likely to experience fever, and the potential selection bias of this sample may have therefore also increased the chances of fever appearing more frequently. It is not possible to tell from our data to what extent one or the other of these possibilities is more likely. Otherwise, the rate of other localized and systemic side effects reported in this sample were similar to that reported in vaccine trials (*40–42*). Awareness of selection bias is important to contextualize survey findings. That said, the sample size for this survey is large enough to suggest that the observed trends are real and are affecting a large number of vaccinated people.

An additional limitation of these analyses is due to the fact that our sample has a very high percentage of people who identify as white and as not Hispanic/Latinx. There are several potential causes for this, including that this data may reflect early trends in vaccination (*84*), it may be related to internet access (though the survey was tested for smartphone functionality), it may be related to participant trust in academic research, and/or it may be a function of how information about the survey was disseminated across social media and traditional media platforms. Social media allows for dissemination, but it also often creates insular communities due to differences in user demographics (*85*). Whatever the cause, we note the under-representation of Black, Indigenous, Latinx, and other respondents of color as a limitation in this research that seeks to understand menstrual experiences after vaccination. One of the ways we have sought to correct this underrepresentation is through the creation of a Spanish language version of the survey, which has only recently concluded.

Overall, our results align with other recent studies which show significant menstrual cycle responsiveness to SARS-CoV-2 vaccination. For example, a Norwegian cohort study found increased reports of heavier periods and longer menstrual bleeding after vaccination which lasted for two to three months (*86*), and U.S.-based sample found longer cycle lengths after vaccination but no effect on bleeding duration (*87*). The only study published thus far which examined menstrual flow after vaccination had similar findings to ours, specifically that that people using hormonal contraceptives were more likely to experience heavier bleeding after vaccination; however, they did not find an association with diagnosed reproductive conditions although they note their sample size might be too small and underpowered for this analysis (*88*). To the best of our knowledge, our work is the first to examine breakthrough bleeding after vaccination in either pre-or post-menopausal people. Furthermore, our large, gender-inclusive sample encompasses a broad age range, allowing us to more closely examine demographic trends and pre-existing health and reproductive factors which narrow down future avenues for further investigation.

Gaps in knowledge of how menstrual cycles respond to acute and chronic immune and inflammatory stressors can be understood as a form of ignorance which is produced and reproduced based upon structural, cultural, and political decisions (*89*). The data presented and discussed here highlights how anthropological mixed-methods research approaches that engage in listening rather than strictly *pro forma* hypothesis-driven research is necessary during emerging phenomena. Taking the time to listen and notice allows us to observe things that may not fit into our established narratives and to take responsibility for our role in knowledge dissemination as scientists (*90, 91*). Furthermore, examination of the narratives and stories that we use to understand the world around us can illuminate the ways scientific narratives can shape and reproduce inequitable power structures of the world. Research which notices and attends to the experiences of people as well as our obligations and relations (*92*) is a necessary first step to building reciprocity (*93*) needed restore trust and create transparency in science.

We have documented a phenotype of increased menstrual bleeding across a diverse set of currently and formerly menstruating people as a post-vaccine response. In doing so, we help provide evidence and context for clinicians regarding the validity of these experiences and we note future avenues of inquiry for researchers. Recognizing and attending to this emerging phenomenon of bleeding changes can help bolster trust between people who menstruate and medical providers, which is an area that has a long history of medical misogyny and gaslighting (*94–97*). Current and historic focus on fertility and reproduction in research and clinical trials is insufficient for addressing the changes in bleeding patterns that cause concern in many people.

We urge other researchers and funding bodies to increase investment in understanding queer, trans, and nonbinary menstrual experiences, because there is a dearth of existing literature to understand the biosocial context of menstrual bleeding in these groups. Furthermore, we note that postmenopausal bleeding remains understudied. Mixed-methods and community based participatory research to address questions that matter to those historically excluded from reproductive and menstruation science is needed in order to provide adequate and culturally and physically relevant care to these populations.

## Materials and Methods

### Recruitment and Survey information

This research was designated as exempt by the University of Illinois Institutional Review Board and Washington University in St. Louis Institutional Review Board. Data were collected and managed using REDCap hosted at the University of Illinois at Urbana-Champaign (*98, 99*). REDCap (Research Electronic Data Capture) is a secure, web-based software platform designed to support data capture for research studies. The survey launched on April 7, 2021, and data for these preliminary results were downloaded on June 29, 2021 (approximately 12 weeks of data collection). The survey was initially announced on Twitter to recruit people who currently or previously menstruated and had been vaccinated (*100, 101*), but it quickly propagated through multiple social media platforms. Media coverage (TV news, public radio, online journalism, print journalism, science blogs, etc.) of the study included links to the survey and provided wide-spread participant recruitment. Additionally, many participants learned of the survey after performing an online search to investigate their own menstrual experiences and finding social media and/or news coverage of this project. Thus, the data collected by this survey represents extensive snowball sampling via many channels.

The survey included a mixture of multiple-choice and text entry questions about typical menstrual experiences (e.g., period flow, cycle length, bleeding duration, common menstrual symptoms), menstrual experiences after each vaccine which make comparison to expected period symptoms (e.g., heavier/lighter/same), other menstrual symptoms, time between vaccine and menstrual side-effects (multiple choice question with ranges), reproductive history (e.g., history of pregnancy, parity, history of post-partum hemorrhage), diagnoses of common reproductive conditions associated with altered menstrual bleeding patterns (e.g., endometriosis, adenomyosis, polycystic ovarian syndrome (PCOS), menorrhagia, fibroids), hormonal treatments (e.g., hormonal contraception, hormonal IUDs, other treatments including gender-affirming hormones such as testosterone), and demographics. Cycle length and number of births were an integer-validated text boxes and most other questions were Yes/No, check boxes, or multiple-choice questions which included an “other” or “not listed here” option to provide a text entry. The survey took approximately 15-20 minutes to complete. Additional details about study variables and the survey can be found in publicly archived supplemental information available at https://osf.io/6rvxk/?view_only=f91f1247658f49e3bbf59b2f6cfd3898.

### Data Cleaning

Data cleaning was performed on select text entries. Specifically, use of gender affirming hormones, reasons for irregular menstruation or non-menstruation, current pregnancy, IUD type, age, and post-vaccine menstrual experience (e.g., breakthrough and menstrual flow) were coded from text responses or from existing survey questions. Common reasons for irregular menstruation or non-menstruation were categorized by combining text entry responses with checkbox options added after the survey was live (i.e., using gender affirming hormones, using long-acting reversible contraceptives, perimenopausal status, postmenopausal status, history of hysterectomy, current or recent lactation, and other). We screened for current pregnancy by evaluating respondent text responses regarding reasons for irregular or non-menstrual status. No respondents in the analyzed sample reported being pregnant. IUD type was determined from text responses and categorized as hormonal, non-hormonal/copper, or unknown type. Respondents with age greater than 99 (N=26) were manually adjusted based on first two numbers entered (e.g., 323 was coded as 32) or by calculating the age from the birth year entered (e.g., 1990 was coded as 31).

Menstrual changes were coded based on survey items across both vaccination doses. Flow change for regularly cycling people was coded based on the dose 1 and dose 2 items assessing period flow with responses lighter, same, or heavier (See Table S9). Due to the proportion of people experiencing heavy flow after at least one of the vaccines, we grouped regularly cycling individuals with any heavier flow into one condition (“heavier”), people who experienced no change in flow after either dose into the second condition (“no change”), and the remainder of people who experienced a combination of lighter and no change after their doses into a smaller third condition (“not heavier”). In total, 727 were missing dose 1 period flow information and 3,031 were missing dose 2 period flow information (Table S9). If information was missing at either dose, we treated it as pairwise missingness, so the respective menstrual change variable was missing. Non-menstruating respondents were categorized as experiencing breakthrough bleeding if they reported spotting, a period, or other menstrual bleeding after either dose.

### Sample

At the time of downloading, 92,529 participants had completed the informed consent and submitted the survey. This included only unique email IDs with duplicate emails being sorted by timestamp and the more recent timestamped responses retained (N=205). Five individuals were removed for inappropriate and/or hostile responses. We removed participants under age 18 (N=12). Responses missing more than 90% of survey items were removed (N=11,999). From the remaining responses (N=80,513), we retained only those that reported having not been diagnosed with COVID-19 (N=65,241; removing N=4,494 with diagnosed COVID-19; 5,761 with suspected but undiagnosed COVID-19; 4,870 who were unsure about prior COVID-19, and 103 reporting “other”) as there is evidence that some people who contract COVID-19 have changes to menstrual bleeding (*34*). There were 42,097 who had received two vaccine doses, 19,161 who had not received a second dose, and 3,983 who did not respond. Two-dose vaccinated individuals were restricted to those who submitted the survey at least 14 days after their second vaccination date (N=35,660). Individuals who received only one dose (N=23,144) were only included if they received the Johnson & Johnson vaccine and completed the survey at least 14 days after first dose vaccination date (N=3,469). In total, we removed 26,112 respondents who were not at least 14 days after full vaccination (i.e., two weeks after second dose for two-dose vaccines or two weeks after vaccination for single-dose vaccines). The final sample was 39,129 participants for general sample descriptive statistics.

We focused on the 35,660 individuals who received a two-dose SARS-CoV-2 vaccination for statistical analyses of menstrual changes and vaccine experiences. Of these respondents, baseline, pre-vaccine menstrual cycles were self-described as regular (N=27,143), irregular (N=4,358), or absent (N=4,136), with 23 individuals not responding to this multiple-choice item and thus excluded from analyses beyond sample description. Analyses focus on conservatively defined subsamples based upon self-reported typical pre-vaccine menstrual cycle status with additional restrictions to reduce the confounding influence of variables that likely affect menstrual cycles. We identified two major groups in the sample– those who regularly menstruate, and those who do not currently menstruate but have in the past. Respondents who regularly menstruate are premenopausal people (ages 18-45) with either spontaneous menstrual cycles or hormonally contracepting cycles who still bleed regularly. Non-menstruating respondents are premenopausal people (ages 18-45) on hormonal treatments that suppress menstruation (e.g., continuous use of hormonal contraceptives, long-acting reversible contraception (LARC), gender-affirming care such as testosterone) and postmenopausal people not on any hormonal treatments (ages 55-80, no period for at least 12 months). The majority of respondents who use gender-affirming care (242 of 267) specified testosterone. We included comparisons to those with diagnosed reproductive conditions generally (e.g., menorrhagia, endometriosis), as well as several specific reproductive conditions hypothesized to be relevant to inflammatory or hemostatic changes in the uterus.

Further details can be found in the Supplement and demographics are reported in Table S4-5. Briefly, we removed respondents who reported having a hysterectomy (N=43), reported currently or recently lactating (N=2,498), and/or gave discrepant responses (e.g., self-reported period details did not align with self-reported menstrual group). The pre-menopausal sample was restricted to age below 45 and post-menopausal sample was restricted above age 55 due to the variability expected throughout perimenopause. People who reported having irregular menstrual cycles or were perimenopausal or at an uncertain menopause stage are not included in these analyses.

### Data Analytic Strategy

We started with descriptive statistics of the full sample of 39,129 fully vaccinated individuals grouped into age categories, omitting all second dose variables for the single-dose Johnson & Johnson vaccine. As this was an emerging phenomenon, we focus primarily on descriptive statistics and trends. We conducted preliminary analyses of associations between menstrual changes (i.e., flow change in menstruating respondents, breakthrough bleeding in non-menstruation respondents) and race, and ethnicity, vaccine type (restricted to the most common 2-dose vaccines, Pfizer and Moderna), vaccine symptoms, typical period experience, reproductive history, and diagnosed reproductive conditions in preliminary univariate analysis of the sample groups using chi-square test of independence and using one-way ANOVA and t-tests for age differences.

We then used a multivariate logistic regression based on the preliminary univariate associations. Specifically, the outcome was whether heavier flow (in regularly menstruation respondents) or breakthrough bleeding (in pre-menopausal non-menstruating respondents) occurred after either dose of the vaccine, and the covariates were vaccine type, race, ethnicity, age, post-vaccine adverse effects of fever and fatigue, diagnosis of a reproductive condition, contraceptive hormone use, history of bleeding during pregnancy, and history of postpartum hemorrhage (PPH). The non-menstruating postmenopausal subgroup was too small for effective use of this statistical approach and thus we present stratified univariate analyses.

As the goal of this paper was to characterize the experiences of a wide range of people, we acknowledge the limitation of significance tests and primarily focus on effect size estimates and odds ratios. However, we also report and incorporate p-values in our analyses and use them in combination with effect size estimates and confidence intervals when discussing results. Our analyses should be considered exploratory and descriptive to aid future hypothesis development to examine menstrual changes experienced following vaccines. All analyses were conducted in R (*102*). DescTools was used for chi-square test power analysis (*103*), rcompanion for Cramer’s *V* (*104*), questionr for odds ratio (*105*), and ggplot2 for figures (*106*). Additional details and supplements for the survey instrument are publicly archived: https://osf.io/6rvxk/?view_only=f91f1247658f49e3bbf59b2f6cfd3898.

## Data Availability

Data is available upon appropriate request in combination with a data use agreement.

## Acknowledgments

We first and foremost would like to thank the tens of thousands of people who responded to the survey to tell us about their experiences. We additionally would like to thank (in alphabetical order) Bryana Rivera, Fatima Soumare, Emma Verstraete, and Florence Yung for their contributions to the project.

## Author contributions

Conceptualization: KL, KC

Methodology: KL, KC, EJ, CL

Investigation: KL, KC

Visualization: EJ, CL, KL, KC

Supervision: KC, KL

Project Administration: KL

Data Curation: KL, EJ

Writing—original draft: KL, KC, EJ, MC Writing—review & editing: KL, KC, EJ, MC, UF, CL

## Data and materials availability

Data is available upon appropriate request in combination with a data use agreement.

## Supplementary Materials for

### Supplementary Text

#### Study Variables

Flow change: The main outcome under investigation for regularly menstruating participants was the changes to period flow during the vaccination time. Two survey items addressed period flow, one for dose 1 and an identical item for dose 2, “After dose 1 [or 2] my period flow was…” with response options “lighter than usual”, “about the same”, or “heavier than usual”. For analysis, changes in bleeding heaviness were coded as “heavier” (based on reports of “heavier than usual” after either dose), “no change” (based on reporting “about the same” after both doses), and a third “heterogeneous no change or lighter” condition (based on a combination of reports of “about the same” or “lighter than usual” after either dose). Those that did not report flow information for either dose was missing flow change variable.

Length change: The second outcome variable for regularly menstruating individuals was change in period length or duration after vaccination. Two identical items measured the change in period length using the stem “After dose 1 [or 2] the length of my period was…” with responses “shorter”, “same”, or “longer”. We coded the following conditions: “longer” (based on reporting “longer” at either dose), “no change” (based on reporting “same” period length after both doses), “heterogeneous no change or shorter” (based on reporting a mixture of seeing no change or a shorter period length), or missing (anyone who failed to report length for either the dose 1 or the dose 2).

Breakthrough experienced: The outcome variable for non-menstruating individuals was the occurrence of breakthrough bleeding (defined as spotting, a period, and/or other menstrual bleeding) after vaccination following vaccine doses. In descriptive reporting, we examined whether breakthrough bleeding occurred after both doses, only dose 1, only dose 2, or did not occur at all. Participants who did not report any breakthrough bleeding are represented in the ‘None’ category. Breakthrough bleeding for statistical analysis was coded as 0= no breakthrough, 1= breakthrough bleeding.

Demographics: The demographic variables used were age, race, and ethnicity. A numerical entry text box asked age. A survey checkbox was used to address race: options were American Indian/Alaskan Native, Asian, Black or African American, Native Hawaiian or Pacific Islander, White, or other. A checkbox was also used for ethnicity with options of Non-Hispanic or Latinx, Hispanic or Latinx, and other. Race was coded as 0= White-only selected, 1=All other selections. Ethnicity was coded as 0=Non-Hispanic/Latinx-only selected, 1=All other selections. We describe all other selections as the diverse racial or diverse ethnic groups due to few participants in each category.

Usual menstrual experiences: Typical period flow and period length were assessed each with one item. Period flow was rated as ‘heavy’, ‘moderate’, ‘light’, ‘non-menstrual’, or ‘other’. Period length, or days with bleeding, were rated as ‘1-3 days long’, ‘3-5 days long’, ‘5-7 days long’, ‘7 or more days long’, ‘non-menstrual’, or ‘other’. The latter categories, non-menstrual and other, were excluded from subgroups as discrepant and from analysis, respectively.

Period symptoms: There were checkboxes to select period symptoms experienced following dose 1 [or 2]. The items were “gotten your period”, “experienced spotting”, “experienced other menstrual bleeding”, “experienced breast/chest soreness”, “experienced symptoms that you typically associate with your period (e.g., cramping, bloating)”, “other”, “I did not have any of these symptoms”.

Timing of period symptoms: Two survey items asked how long following dose 1 [or 2] before participants experienced the reported period symptoms. The options were 1-3 days, 4-7 days, 8-14 days, more than 14 days, or cycling at the time of the dose.

Vaccine symptoms: Participants responded to a single item asking whether they experienced any side effects following the first dose and, similarly, for the second dose. Subsequent items for vaccine symptoms following the first dose and second dose included checkboxes for arm-soreness, fever, fatigue, headache, nausea, or other. Fever and fatigue were identified as meaningful to compare alongside bleeding conditions, because they are more unrelated to a normal menstrual experience than headaches, for example. Fever was coded as 1 if participants said yes following either dose and 0 if it was not checked after either dose. Fatigue was, similarly, coded as fatigue following either or no fatigue after either dose.

Reproductive history: Participants answered whether they had ever been pregnant (coded 1=pregnancy, 0=no). They were asked, also, if they had ever given birth (1=parous, 0=no). A history of bleeding at either event were captured with the items, “Did you experience any vaginal bleeding during your pregnancies” and “Did you experience postpartum hemorrhage with any of your births”. Each subsequent question appeared contingent on having experienced the event of pregnancy or birth.

Reproductive conditions: A checkbox item assessed, “Have you ever been diagnosed with any of the following” with options heavy menstrual bleeding, abnormal uterine bleeding, menorrhagia, endometriosis, adenomyosis, fibroids, polycystic ovarian syndrome, and/or other condition you feel is relevant. The first three options were common descriptions given to describe a similar condition, which we refer singularly to as menorrhagia.

### Additional details for subgroup definitions

#### 1. Subgroups of Menstruating Sample

The first group identified in the full sample were the individuals who reported not being diagnosed with any reproductive conditions (i.e., PCOS, endometriosis, menorrhagia or similar bleeding disorders, etc.), which likely affect people’s menstrual experience. People reported their typical menstrual cycle as regularly occurring (typically 20-40 day cycles that feel predictable), irregular or occasional (very far apart, not predictable, or both), or rarely or do not menstruate right now.

##### 1.1. Regularly Cycling Individuals

Those that reported regularly menstruating were restricted to a conservative group between the ages of 18 and 45 years-old, were not lactating in the last year, and had no history of hysterectomy.

###### 1.1.1. Spontaneous Regular Cyclers

The conservative sample for regularly menstruating individuals that were not on any hormones (including birth control, thyroid treatment, other hormones) was 12,364. Discrepant responders were removed based on describing their usual period flow as ‘non-menstrual’ (n=4). Then we removed individuals that responded as not having a period after both doses (n=660). This subgroup of spontaneous regularly cycling comprised 11,700 participants. In the sample, 780 individuals reported having a copper, or non-hormonal IUD.

###### 1.1.2. Hormonally Contracepting Regular Cyclers

The conservative sample for regularly menstruating individuals that were on hormonal contraceptives and/or other hormones was 4,185. Discrepant responders were removed based on describing their usual period flow or period length as ‘non-menstrual’ (n=25). Then we removed individuals that responded as not having a period after both doses (n=305). The subgroup comprised 3,855 participants that are hormonally contracepting and regularly cycling.

##### 1.2. Non-menstruating Individuals

Those that reported not menstruating for various reasons included 673 postmenopausal, 280 on gender affirming hormones/undergoing gender affirmative therapy, 1,911 on Long-Acting Reversible Contraceptives (LARC), 48 had hysterectomies (full or partial), 274 coded as being in an uncertain menopause stage, 463 peri-menopausal, 329 lactating recently, and 321 selected other responses not listed. Multiple reasons were able to be selected. We removed those that reported lactating recently (n=) or had a history of hysterectomy (n=48).

###### 1.2.1. Individuals undergoing Gender Affirmative Care

The conservative sample of non-menstruating cycling individuals who described masculinizing therapy (e.g., testosterone) or reported undergoing gender affirming care was restricted to ages 18 to 45 years-old and, by definition, on hormones. Due to the use of some forms of birth control for gender affirmative treatment, we instead describe the categories of hormones the sample reported: 9 hormonal contraception only, 137 other hormones (e.g., testosterone) only, or 37 a mixture of both. In total, 27 were on hormonal contraceptives (i.e., birth control), 20 hormonal IUDs, 9 non-hormonal (copper) IUDs, and 174 reported other hormones. The gender-affirmative subgroup comprised 183 individuals.

###### 1.2.2. Individuals on Long-Acting Reversible Contraceptives

The conservative sample of those on Long-acting Reversible Contraceptives (LARC) and non-menstruating cycling were restricted to ages between 18 and 45 years-old and, by definition, were on hormones. There are 41 individuals who are, also, represented in the gender affirmative group, so we removed them from the LARC subgroup. The hormones participants reported were grouped as 914 hormonal contraception only and 29 reported a mixture of other hormones and hormonal contraception. In total, 280 on hormonal contraceptives (i.e., birth control), 684 hormonal IUDs, 2 non-hormonal IUDs, and 29 on other hormonal treatments. The LARC subgroup was 943 individuals.

#### 2. Subgroups of Menstruating Sample diagnosed with Reproductive Conditions

The second group identified in the full sample were individuals that were diagnosed with one of a number of reproductive conditions (i.e., menorrhagia, endometriosis, adenomyosis, fibroids, PCOS, PPH, or other). There were 11,502 in the full sample that reported being diagnosed with at least one condition. The mean age was 36.28 years-old (SD=9.45 years; range from 18 to 77). Of this group, 7,774 were regularly menstruating, 1,949 were irregular, 1,768 were non-menstruating, and 11 did not respond. Two of the irregularly cycling participants reported having a full or partial hysterectomy along with 45 of the non-menstruating. So, we removed the individuals who reported having had a hysterectomy and those who were lactating recently, post-menopause, perimenopause, or in an uncertain menopause stage (n=10,105). Finally, we restricted the ages to 18 to 45 years-old in a conservative look at the menstrual changes to individuals diagnosed with reproductive conditions. The final sample number was 8,652.

##### 2.1.1. Individuals that are Spontaneous Regular Cyclers

From the final reproductive conditions sample we first partitioned out individuals described as regularly cycling spontaneously. This subgroup included 4,013, which was further reduced by removing discrepant responses of ‘non-menstrual’ to either normal period flow and length (n=7). Anyone who reported not having had a period after both dose 1 and dose 2 were removed (n=221). Therefore, this spontaneous regularly cycling subgroup comprised N=3,785 participants. Non-hormonal IUDs were reported for 201 individuals in the subgroup.

##### 2.1.2. Hormonally Contracepting Individuals that are Regular Cyclers

From the regularly cycling individuals in the reproductive conditions sample, we next focused on those hormonally contracepting (including those on other hormonal treatments or medications). This subgroup consisted of 2,218 individuals before we removed the discrepant responses (n=13) and those not having had a period (n=165). The final subgroup included 2,040 participants.

##### 2.1.3. Hormonally Contracepting Individuals that are Non-menstruating

The portion of the sample diagnosed with a reproductive condition and described themselves as not menstruating was 942 individuals. Only 68 individuals were not on any hormonal treatments or medications, so we focused on hormonally contracepting individuals aged 18 to 45 diagnosed with at least one reproductive condition and described themselves as non-menstruating. The subgroup of non-menstruating individuals was 874. The majority of the sample described themselves as on LARC. And a small portion describes themselves as using gender-affirming hormones. Therefore, we focus on exclusive subgroups of LARC individuals and people on gender-affirming hormones.

###### 2.1.3.1. Individuals Undergoing Gender Affirmative Care

The hormones reported by this group were 4 on hormonal contraceptives only, 57 on hormonal treatments only, and 26 on mixture of both: more specifically, 17 hormonal contraceptives, 14 hormonal IUDs, 2 copper IUDs, and 83 on other hormonal treatments. The final sample of people diagnosed with a reproductive condition and non-menstruating undergoing gender-affirmative care were 87 individuals.

###### 2.1.3.2. Individuals on Long-Acting Reversible Contraceptives

There are 24 individuals who are represented in the gender affirmative group, also, represented in the LARC group, so we removed them from LARC subgroup. The categories of hormones used were 549 on contraceptive hormones only, 2 on other hormones only (progestin), and 51 mixture of both: more specifically, 209 on hormonal contraceptives, 410 hormonal IUDs, 2 copper IUDs, and 53 on other hormonal treatments. The final sample for people diagnosed with a reproductive condition and non-menstruating on LARC were 602 individuals.

##### 2.1.4. Subgroups of Post-menopause Sample

###### 2.1.4.1. Post-menopause without diagnosed reproductive conditions

The conservative sample for post-menopause individuals was restricted to ages 55 or older. Due to small sample numbers, we restricted to those not on any hormones (including birth control, thyroid treatment, other hormones). The subgroup post-menopause consisted of 117 individuals.

###### 2.1.4.2. Post-menopause with diagnosed reproductive conditions

The conservative sample for post-menopause individuals who have a diagnosed reproductive condition was restricted to ages 55 or older and not on any hormones. The subgroup of post-menopause individuals with a diagnosed condition was 121 individuals.

**Table S1. Full reporting of demographics and sample background. (separate file)**

Expanded demographics and sample background. Full reporting for in-text Table 1.

**Table S2. Menstrual changes and vaccine symptoms reported after each vaccine dose (full reporting). (separate file)**

The reported symptoms and changes are grouped by age and by vaccine type. Dose 1 and dose 2 sample sizes differ because the Johnson&Johnson vaccine does not have a second dose, so participants do not report on any dose 2 survey items. Dose 1 and 2 period flow were used to calculate flow changes in regularly menstruating age groups. Period symptoms (had a period, spotting, and other menstrual bleeding) were used to calculate whether breakthrough occurred in non-menstruating age groups.

**Table S3. Menstrual changes and vaccine symptoms reported after each vaccine dose (abbreviated reporting). (separate file)**

The reported symptoms and changes as reported in Table S2 are grouped by age but not by vaccine type.

**Table S4. Subsample demographics and background. (separate file)**

The demographic reporting of the regularly cycling premenopausal respondents, the premenopausal non-menstruating respondents, and the postmenopausal respondents.

**Table S5. Menstrual changes and vaccine symptoms reported after each vaccine dose. (separate file)**

The reported symptoms and changes are displayed for the three sample groups: premenopausal regularly cycling, premenopausal non-menstruating, and postmenopausal respondents.

**Table S6. Menstrual and medical history related to changes in flow across regularly cycling subgroups. (separate file)**

These results refer to the univariate analyses of regularly cycling respondents for associations with flow change. Four subgroups are examined: spontaneous cycling with diagnosed reproductive conditions, the same without diagnosed reproductive conditions, hormonally contracepting with diagnosed reproductive conditions, and the same without diagnosed reproductive conditions.

**Table S7. Proportion comparisons between premenopausal regular-cycling reproductive condition diagnoses and those without diagnosed reproductive conditions. (separate file)**

These results refer to the univariate tests for associations between specific reproductive conditions versus no diagnosed condition and flow change.

**Table S8. Vaccine and medical history related to breakthrough bleeding across pre-menopause non-menstruating groups. (separate file)**

These results refer to the univariate analyses of premenopausal non-menstruating respondents for associations with breakthrough bleeding. The two groups are examined: those on LARC and those on gender affirming hormones.

**Table S9. Contingency table of menstrual changes in all regularly cycling individuals (N=21**,**380)**.

The dose 1 period flow and dose 2 period flow contingency table displayed for all regularly cycling individuals.

